# Prediction Model for Severe Community-acquired Pneumonia Development among Patients with Diabetes Mellitus

**DOI:** 10.1101/2021.02.04.21251184

**Authors:** Ruoming Tan, Tingting Pan, Yuzhen Qiu, Jiahui Wang, Xiaoling Qi, Erzhen Chen, Min Zhou, Jialin Liu, Hongping Qu

## Abstract

**Introduction:** Diabetes is an independent risk factor for the development of severe community-acquired pneumonia (CAP) and associated with pneumonia-related hospitalization as well as mortality. Here, we assessed several selected biomarkers to determine their predictive value for progression to severe CAP among diabetic patients.

**Research design and methods:** A retrospective cohort study of diabetic patients with CAP was conducted at a tertiary teaching hospital. The prediction model group (N=100) comprised patients registered between January 2015 to December 2016. Multivariate analysis was performed to identify predictive biomarkers from this cohort. The validation group (N=108) comprised the patients between January 2017 to February 2019. Predictive performance was assessed in the validation group.

**Results:** A total of 208 diabetic inpatients with CAP were recruited. Further multivariate analysis showed that C-reactive protein (CRP), absolute lymphocyte count (ALC), immunoglobulin (IgM), and HbA1C at admission were independently associated with progression to severe CAP in diabetic patients during hospitalization. The prediction model =0.0179555*CRP+1.975918* HbA1C-2.879364* ALC −0.026255* IgM − 8.220555. The area under ROC (AUROC) curve in the validation group was 0.851 (0.781–0.921) with statistical significance (P<.05).

**Conclusions:** In diabetic patients with CAP, a combination of CRP, ALC, IgM, and HbA1C at admission could be used to predict the progression to severe CAP.

Significance of this study

What is already known about this subject?

1. Diabetes is an independent risk factor for the development of severe community-acquired pneumonia (CAP) and associated with pneumonia-related hospitalization as well as mortality.
2. It is essential to identify these patients early and prevent them from progressing to SCAP to improve clinical outcomes.

What are the new findings?

1. C-reactive protein (CRP), absolute lymphocyte count (ALC), immunoglobulin (IgM), and HbA1C at admission were independently associated with progression to severe CAP in diabetic patients during hospitalization.
2. The prediction model = 0.0179555*CRP+1.975918* HbA1C-2.879364* ALC −0.026255* IgM − 8.220555.
3. The area under ROC (AUROC) curve in the validation group was 0.851 (0.781–0.921) with statistical significance (P<.05).

How might these results change the focus of research or clinical practice?

1. In patients with diabetes and CAP, HbA1C, CRP, ALC, IgM at admission, and their combination could be used to predict the progression to severe CAP during hospitalization with good accuracy.
2. Due to the included biomarkers are common, our finding may be performed well in clinical practice and then improve the early management of diabetic patients with CAP.

## Introduction

Diabetes mellitus is a common chronic disease with multiple complications and contributes to the global health care burden. It is estimated that in 2011, 366 million patients with diabetes were reported worldwide, and by 2030, the number of patients with diabetes will increase by 50% ^1^. In addition, in the United States, diabetes is reported as one of the ten leading causes of death.

Infection is one of the severe complications in patients with diabetes, and the incidence varied between 32.7% and 90.5% ^2,3^. The lower respiratory tract is thought as the most common site involved with infection in diabetic patients ^4–6^. Due to the immunocompromised state, patients with diabetes appear more susceptible to pneumonia than patients without diabetes. Besides this, these patients have an increased risk of hyperglycemia, impaired lung function, and other chronic complications (such as heart disease, renal failure, and pulmonary microangiopathy) ^7–9^. In recent studies, it was demonstrated that diabetes is an independent risk factor for the development of severe community-acquired pneumonia (CAP) and associated with pneumonia-related hospitalization as well as mortality ^8, 10–14^. Hence, based on the above mentioned, to develop a predictive tool for the progression of CAP to severe CAP in patients with diabetes may be useful to improve the management of diabetic patients with CAP.

Unfortunately, limited tools were available for the prediction of severe CAP among diabetic patients with CAP. In the study, the risk factors for severe CAP among patients with CAP were evaluated, then selected variables, and their combination was assessed for predicting the presence of severe CAP among diabetic patients with CAP.

## Methods

### Design and setting

A retrospective cohort study was conducted at a tertiary teaching hospital between January 2015 to February 2019. The study protocol was approved by the Ruijin Hospital Ethics Committee, Shanghai Jiaotong University School of Medicine, China. Informed consent from study patients was not required because of the retrospective design of the cohort study.

### Patients selection and definitions

A total of 208 diabetic patients hospitalized with CAP were recruited. We divided the patients according to their year of pneumonia diagnosis. The prediction model group (N=100) comprised patients registered between January 2015 to December 2016, and the validation group (N=108) included the patients between January 2017 to February 2019. The basic characteristics of the two cohorts are listed in Table 1. CAP was defined by the following criteria: 1) acute lower respiratory tract infection with at least two symptoms (e.g., fever, cough or sputum production, dyspnea, chest pain); 2) new focal signs on physical examination of the chest; 3) new infiltrates on chest x-ray ^15^.

**Table 1.**
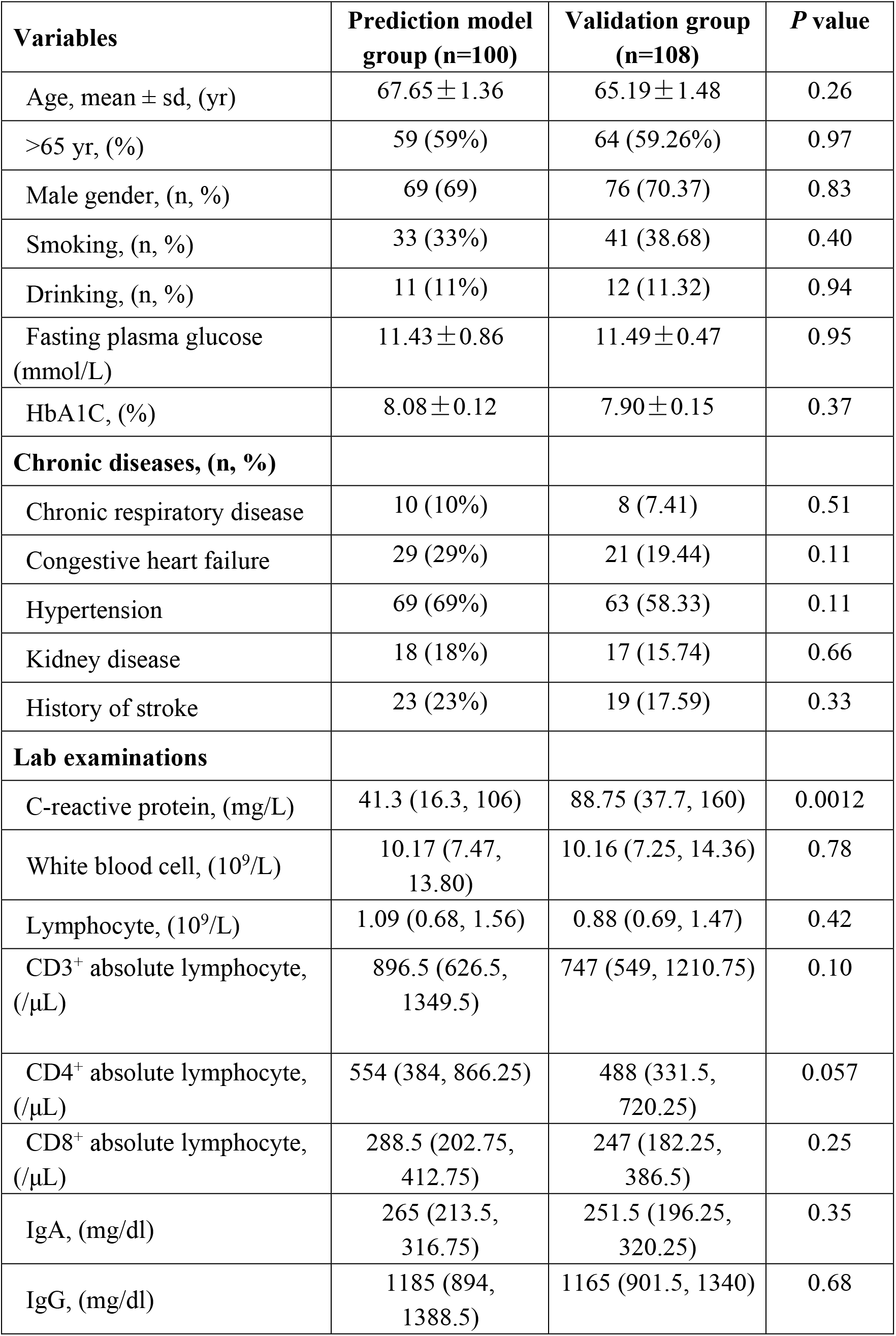
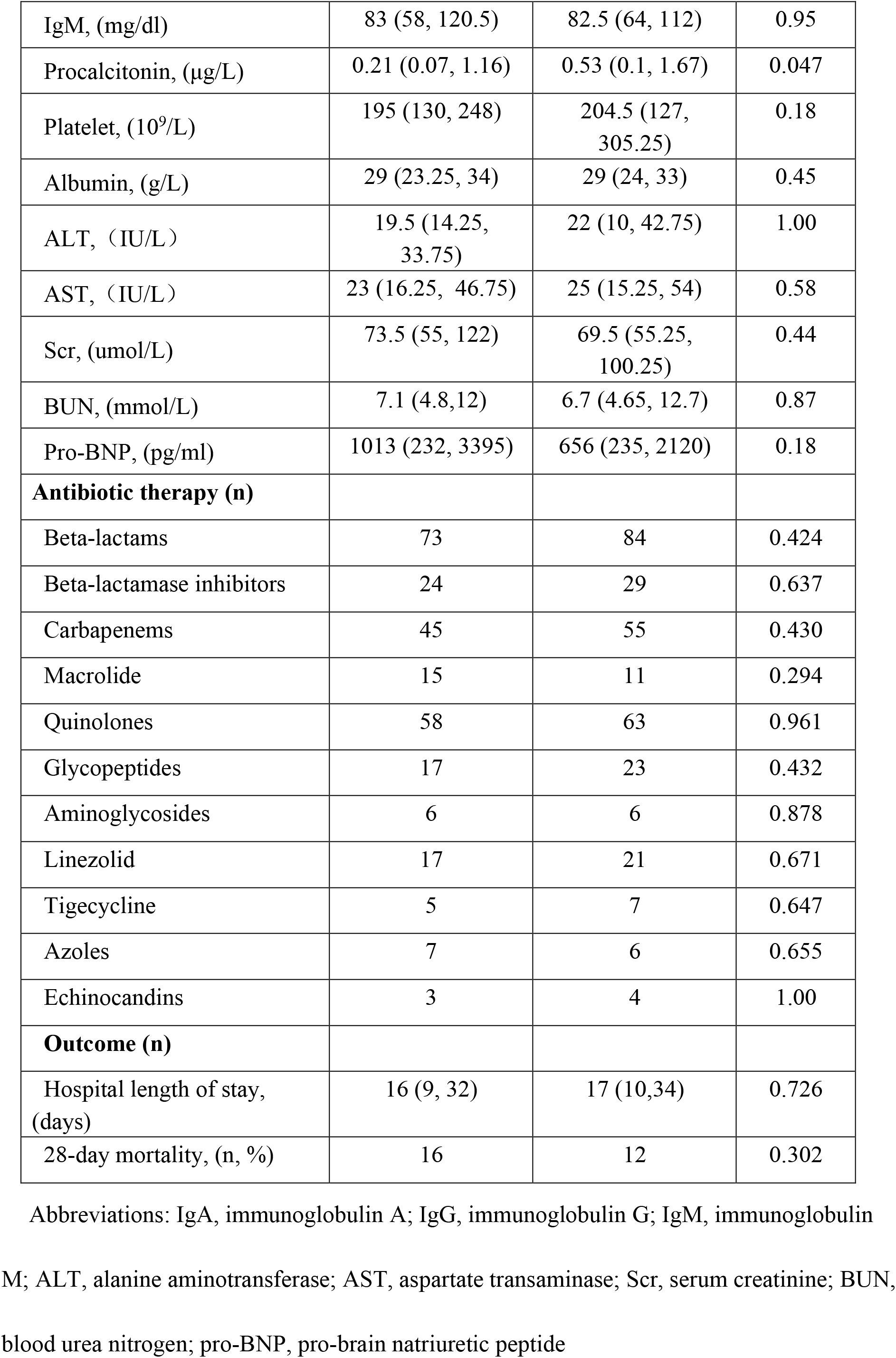
Baseline characteristics of DM patients in prediction model group and validation group.

| Variables | Prediction model group (n=100) | Validation group (n=108) | *P* value |
| --- | --- | --- | --- |
| Age, mean ± sd, (yr) | 67.65 ± 1.36 | 65.19 ± 1.48 | 0.26 |
| >65 yr, (%) | 59 (59%) | 64 (59.26%) | 0.97 |
| Male gender, (n, %) | 69 (69) | 76 (70.37) | 0.83 |
| Smoking, (n, %) | 33 (33%) | 41 (38.68) | 0.40 |
| Drinking, (n, %) | 11 (11%) | 12 (11.32) | 0.94 |
| Fasting plasma glucose (mmol/L) | 11.43 ± 0.86 | 11.49 ± 0.47 | 0.95 |
| HbA1C, (%) | 8.08 ± 0.12 | 7.90 ± 0.15 | 0.37 |
| **Chronic diseases, (n, %)** |  |  |  |
| Chronic respiratory disease | 10 (10%) | 8 (7.41) | 0.51 |
| Congestive heart failure | 29 (29%) | 21 (19.44) | 0.11 |
| Hypertension | 69 (69%) | 63 (58.33) | 0.11 |
| Kidney disease | 18 (18%) | 17 (15.74) | 0.66 |
| History of stroke | 23 (23%) | 19 (17.59) | 0.33 |
| **Lab examinations** |  |  |  |
| C-reactive protein, (mg/L) | 41.3 (16.3, 106) | 88.75 (37.7, 160) | 0.0012 |
| White blood cell, ($10^9$/L) | 10.17 (7.47, 13.80) | 10.16 (7.25, 14.36) | 0.78 |
| Lymphocyte, ($10^9$/L) | 1.09 (0.68, 1.56) | 0.88 (0.69, 1.47) | 0.42 |
| CD3<sup>+</sup> absolute lymphocyte, (/μL) | 896.5 (626.5, 1349.5) | 747 (549, 1210.75) | 0.10 |
| CD4<sup>+</sup> absolute lymphocyte, (/μL) | 554 (384, 866.25) | 488 (331.5, 720.25) | 0.057 |
| CD8<sup>+</sup> absolute lymphocyte, (/μL) | 288.5 (202.75, 412.75) | 247 (182.25, 386.5) | 0.25 |
| IgA, (mg/dl) | 265 (213.5, 316.75) | 251.5 (196.25, 320.25) | 0.35 |
| IgG, (mg/dl) | 1185 (894, 1388.5) | 1165 (901.5, 1340) | 0.68 |
| IgM, (mg/dl) | 83 (58, 120.5) | 82.5 (64, 112) | 0.95 |
| Procalcitonin, (µg/L) | 0.21 (0.07, 1.16) | 0.53 (0.1, 1.67) | 0.047 |
| Platelet, (10<sup>9</sup>/L) | 195 (130, 248) | 204.5 (127, 305.25) | 0.18 |
| Albumin, (g/L) | 29 (23.25, 34) | 29 (24, 33) | 0.45 |
| ALT, (IU/L) | 19.5 (14.25, 33.75) | 22 (10, 42.75) | 1.00 |
| AST, (IU/L) | 23 (16.25, 46.75) | 25 (15.25, 54) | 0.58 |
| Scr, (umol/L) | 73.5 (55, 122) | 69.5 (55.25, 100.25) | 0.44 |
| BUN, (mmol/L) | 7.1 (4.8,12) | 6.7 (4.65, 12.7) | 0.87 |
| Pro-BNP, (pg/ml) | 1013 (232, 3395) | 656 (235, 2120) | 0.18 |
| **Antibiotic therapy (n)** |  |  |  |
| Beta-lactams | 73 | 84 | 0.424 |
| Beta-lactamase inhibitors | 24 | 29 | 0.637 |
| Carbapenems | 45 | 55 | 0.430 |
| Macrolide | 15 | 11 | 0.294 |
| Quinolones | 58 | 63 | 0.961 |
| Glycopeptides | 17 | 23 | 0.432 |
| Aminoglycosides | 6 | 6 | 0.878 |
| Linezolid | 17 | 21 | 0.671 |
| Tigecycline | 5 | 7 | 0.647 |
| Azoles | 7 | 6 | 0.655 |
| Echinocandins | 3 | 4 | 1.00 |
| **Outcome (n)** |  |  |  |
| Hospital length of stay, (days) | 16 (9, 32) | 17 (10,34) | 0.726 |
| 28-day mortality, (n, %) | 16 | 12 | 0.302 |

Abbreviations: IgA, immunoglobulin A; IgG, immunoglobulin G; IgM, immunoglobulin M; ALT, alanine aminotransferase; AST, aspartate transaminase; Scr, serum creatinine; BUN, blood urea nitrogen; pro-BNP, pro-brain natriuretic peptide

Diabetes mellitus was considered to be present if diagnosed in the past medical history. Severe CAP was diagnosed according to the guidelines issued by Infectious Diseases Society of America/American Thoracic Society (IDSA/ATS) and defined if 1) met one of the major criteria: acute respiratory failure requiring invasive mechanical ventilation, septic shock with the need for vasopressors; 2) met at least three minor criteria: respiratory rate ≥30 bpm, a ratio of partial pressure of arterial oxygen to fraction of inspired oxygen (PaO2/FiO2) ≤250, blood urea nitrogen (BUN) ≥20 mg/dL, white blood cell count <0.4×10^9^/L, platelet count <100×10^9^/L, body temperature <36 °C, multi-lobar infiltrates, confusion/disorientation, and hypotension requiring aggressive fluid resuscitation ^15^.

Exclusion criteria were as follows: age less than 18 years, treatment with polyclonal intravenous immunoglobulins in the previous three months, an immunocompromised condition, or insulin requirement during the hospital stay without a prior history of diabetes. Patients with a lack of cellular and humoral immunity analysis data were also excluded. Immunosuppression was defined as oncology; hematology; human immunodeficiency virus infection; treatment ≥ 20 mg corticosteroids daily for ≥14 days; chemotherapy or radiation therapy during the previous six months; immunosuppressive therapy after organ or bone marrow transplantation; and active tuberculosis ^16.^

### Data collection

We collected data on demographic characteristics, symptoms and vital signs, comorbidities (chronic respiratory disease, heart, kidney, liver, and neurologic disorders), and the details of treatment for each patient during hospitalization, such as mechanical ventilation and renal replacement. Laboratory data of blood cell counting, hepatic, renal, cardiac function test, cellular and humoral immunity analysis during the first 72 hours after the diagnosis of CAP were recorded.

### Lymphocyte subset and immunoglobulin analysis

Samples of peripheral blood were collected in ethylenediaminetetraacetic acid tubes during the first 72 hours after CAP diagnosis. This measurement was carried out by using the automatic analyzers available at the central laboratory, in accordance with approved standard operative procedures for clinical use.

### Statistical analysis

The data are reported as No. (%) or as median (interquartile range [IQR]) for categorical and continuous variables, respectively. All variables were tested for normal distribution using the Kolmogorov-Smirnov test. The Student *t*-test was used to compare the means of continuous variables and the normality of data distribution. Otherwise, the Mann-Whitney U-test was used. Categorical data were tested using the chi-square (χ2) test. Of the clinical baseline characteristics, only those with *P* values < .05 by univariate analysis were candidates for the multivariate model, which was finally determined with a backward stepwise variable selection procedure. The multivariate logistic regression analyses were summarized by estimating the odds ratios (OR) and respective 95% confidence intervals (CI). To evaluate the predictive value of the combination of new predictors to predict severe CAP occurrence, we constructed a predictive logistic regression model. Predictive performance was assessed in the evaluation group by estimating negative and positive predictive values, sensitivity and specificity, and the AUC with receiver operating-characteristic (ROC) curves, with corresponding 95% CI. All *P* values were two-sided, with values < 0.05 considered statistically significant. Statistical analyses were performed using Stata 12.0 for Windows (StataCorp, TX, USA).

## Results

### Patient clinical characteristics data

A total of 208 CAP patients with DM (64 women and 144 men) were included, and the patients were divided into the prediction model group (N=100) registered between January 2015 to December 2016 and the validation group (N=108) between January 2017 to February 2019. The mean age of these patients at admission was 69 years old (95% CI, 57 to 77.75). 109 patients progressed to severe pneumonia, thus classified into severe pneumonia-, and 99 patients into non-severe pneumonia groups. The most common underlying condition for all participants was cardiovascular disease (24%). The mean length of stay of the patients with CAP was 16 days, and 16 (7.7%) died within 28 days after diagnosis. The demographic and clinical characteristics of the prediction model group and the validation group are shown in Table 1.

### Comparison of the indicators between DM patients with severe CAP and non-severe CAP in prediction model group

As shown in Table 2, the diabetic patients who developed severe CAP showed higher levels of fasting plasma glucose and HbA1C as compared with non-severe CAP patients. Besides, of the patients with severe CAP, the incidences of comorbid respiratory and renal disease were increased than those in non-severe CAP patients.

**Table 2.** Comparison of the indicators between DM patients with severe CAP and non-severe CAP in the prediction model group.

| **Variables** | **Severe pneumonia**<br>**(n=49)** | **Non-severe pneumonia**<br>**(n=51)** | ***P* value** |
| --- | --- | --- | --- |
| Age mean ± SD (yrs) | 67.96 ± 1.81 | 67.35 ± 2.03 | 0.82 |
| >65 yrs (%) | 68 (62.39%) | 55 (55.56%) | 0.32 |
| Male gender (n %) | 35 (71.43%) | 34(66.67%) | 0.61 |
| Smoking (n %) | 14 (28.57) | 19 (37.25) | 0.36 |
| Drinking (n %) | 5 (10.20) | 6 (11.76) | 0.80 |
| Fasting plasma glucose (mmol/L) | 10.43 (8.81, 13.11) | 9.04 (7.25, 11.72) | 0.036 |
| HbA1C (%) | 8.5 (7.8,9.15) | 7.4 (7.2,7.8) | <0.0001 |

| **Chronic diseases, (n, %)** |  |  |  |
| --- | --- | --- | --- |
| Chronic respiratory disease | 2 (4.08%) | 8 (15.69%) | 0.053 |
| Congestive heart failure | 17 (34.69%) | 12 (23.53%) | 0.22 |
| Hypertension | 36 (73.47) | 33 (64.71) | 0.34 |
| Kidney disease | 15 (30.61%) | 3 (5.88%) | 0.001 |
| History of stroke | 12 (24.49%) | 11 (21.57%) | 0.73 |
| **Lab examinations** |  |  |  |
| C-reactive protein (mg/L) | 79.1<br>(28.55,139.25) | 22 (7.4,64) | <0.0001 |
| White blood cell (10<sup>9</sup>/L) | 12.19 (8.67,<br>14.70) | 8.94 (7.23,<br>11.52) | 0.0031 |
| Lymphocyte (10<sup>9</sup>/L) | 0.8 (0.56, 1.14) | 1.4(0.95,1.91) | <0.0001 |
| CD3<sup>+</sup> absolute lymphocyte (/μL) | 678 (489,911.5) | 1259<br>(888,1588) | <0.0001 |
| CD4<sup>+</sup> absolute lymphocyte (/μL) | 433 (286.5, 538) | 799 (554,<br>1088) | <0.0001 |
| CD8<sup>+</sup> absolute lymphocyte (/μL) | 234<br>(120.5,303.5) | 390 (255, 531) | <0.0001 |
| IgA (mg/dl) | 234 (190,320) | 284 (249, 298) | 0.057 |
| IgG (mg/dl) | 980 (853, 1190) | 1300<br>(1180,1476) | <0.0001 |
| IgM (mg/dl) | 66 (47.5,78.5) | 108 (88,131) | <0.0001 |
| Procalcitonin (μg/L) | 0.7 (0.24,2.08) | 0.09<br>(0.05,0.19) | <0.0001 |
| Platelet (10<sup>9</sup>/L) | 151 (99,226) | 216.5<br>(179.75,264.5) | 0.0005 |
| Albumin (g/L) | 26 (21.5,31) | 32 (27,37) | <0.0001 |
| ALT (IU/L) | 21 (14 ,37) | 19 (15,31) | 0.77 |
| AST (IU/L) | 35 (18.5,55) | 20 (16, 31) | 0.0055 |
| Scr (umol/L) | 97 (53.5,186.5) | 70 (55,88) | 0.037 |
| BUN (mmol/L) | 10.35<br>(7.025,15.05) | 5.5 (3.8,7.8) | <0.0001 |
| Pro-BNP (pg/ml) | 2198 (606,<br>8698.5) | 571.3 (92.625,<br>1476.75) | 0.0001 |

| Microbiological findings (n) |  |  |  |
| --- | --- | --- | --- |
| Gram-positive | 4 | 0 | 0.054 |
| Gram-negative | 5 | 2 | 0.264 |
| Fungi | 1 | 4 | 0.264 |
| Virus | 4 | 2 | 0.432 |
| Unknown | 38 | 44 | 0.256 |
| Mixed | 3 | 1 | 0.357 |

Abbreviations: IgA, immunoglobulin A; IgG, immunoglobulin G; IgM, immunoglobulin M; ALT, alanine aminotransferase; AST, aspartate transaminase; Scr, serum creatinine; BUN, blood urea nitrogen; pro-BNP, pro-brain natriuretic peptide

Compared to DM patients with non-severe CAP, severe CAP patients showed significantly higher values for the CRP, WBC, PCT, AST, Scr, BUN, and pro-BNP (Table 2) that DM patients with severe CAP may develop more complications.

Of note, the lymphocyte subsets counts were analyzed by the CAP severity (Table 3). DM patients with severe CAP had lower ALCs and lower CD3^+^, CD4^+^, and CD8^+^ T cell counts than those in non-severe CAP patients. In addition, DM patients with severe CAP showed significantly lower values for the IgM and IgG (Table 2). Serum immunoglobulin levels decreased in relation to the severity of CAP in DM patients.

**Table 3.** Univariate and multivariate logistic regression analysis of risk factors for progression to severe pneumonia among DM patients with CAP.

|  | Univariate logistic regression |  |  | Multivariate logistic regression |  |
| --- | --- | --- | --- | --- | --- |
| Variables | OR (95% CI) | *p* valve | Coefficient | OR (95% CI) | *p* valve |
| C-reactive protein mg/L | 1.013(1.005,1.020) | 0.0001 | 0.013 | 1.018(1.003,1.033) | 0.017 |
| **Kidney disease** | 8.82 (2.38, 32.67) | 0.0002 | 2.177 | 4.438(0.152, 129.19) | 0.386 |
| **Hypertension** | 1.51( 0.64, 3.55) | 0.343 | 0.412 |  |  |
| FBG | 1.000(0.955, 1.047) | 0.996 | 0.0001 |  |  |
| HbA1C | 4.04(2.13, 7.67) | <0.0001 | 1.40 | 7.213(1.769, 20.41) | 0.006 |
| White blood cell 10<sup>3</sup>/mm<sup>3</sup> | 1.13(1.02,1.24) | 0.01 | 0.122 | 1.132(0.938,1.366) | 0.197 |
| lymphocyte | 0.173(0.073,0.408) | <0.0001 | -1.76 | 0.056(0.0067,0.471) | 0.008 |
| IgG (mg/dl) | 0.999(0.997,1.000) | 0.010 | -0.0015 | 0.999 (0.997,1.002) | 0.564 |
| IgM (mg/dl) | 0.979(0.968,0.991) | <0.0001 | -0.021 | 0.974(0.951,0.998) | 0.033 |
| Procalcitonin µg/L | 1.117(0.966,1.291) | 0.017 | 0.111 | 1.343(0.936,1.928) | 0.109 |
| Platelet 10<sup>3</sup>/mm<sup>3</sup> | 0.994(0.989,0.998) | 0.005 | -0.00628 | 0.995(0.987,1.004) | 0.325 |
| Albumin (g/L) | 0.862(0.799,0.930) | <0.0001 | -0.148 | - | - |
| AST (IU/L) | 1.032(1.010,1.054) | <0.0001 | 0.0316 | 0.892(0.768,1.036) | 0.136 |
| Scr (umol/L) | 1.007(1.0015,1.0128) | 0.0016 | 0.0070 | 1.022(0.979,1.067) | 0.325 |
| BUN (mmol/L) | 1.174(1.067,1.291) | <0.0001 | 0.160 | 0.998(0.984,1.011) | 0.728 |
| Pro-BNP (pg/ml) | 1.000 (1.000,1.000) | <0.0001 | 0.00032 | 0.995(0.967,1.025) | 0.755 |

Abbreviations: FBG, fasting blood glucose; IgG, immunoglobulin G; IgM, immunoglobulin
M; AST, aspartate transaminase; Scr, serum creatinine; BUN, blood urea nitrogen; pro-BNP, pro-brain natriuretic peptide

### Analysis of the risk factors for progression to severe pneumonia in DM patients with pneumonia admitted

To determine early prediction factors of severe pneumonia development in DM patients with pneumonia admitted, we performed a univariate logistic regression analysis. The results showed that CRP, kidney disease, HbA1C, ALC, IgM, IgG, PCT,PLT, albumin, AST, Scr, BUN, and pro-BNP might be used as predictive factors for severe pneumonia development in DM patients with pneumonia admitted.

To further determine the independent predictive factors for severe pneumonia development in DM patients, multivariate logistic regression analysis was performed. In multivariate analysis, CRP, HbA1C, ALC, and IgM were independently associated with severe CAP development in DM patients (Table 3).

### Development of a prediction model and validating in the validation group

We developed a prediction model to determine the weight of independent prediction factors of severe pneumonia development in DM patients. The prediction model was established based on multivariate analysis: the prediction model =0.0179555*CRP+1.975918* HbA1C-2.879364* ALC −0.026255* IgM − 8.220555.

The validation group was used to further validate the predictive accuracy of this model. Different threshold performances including sensitivity, specificity, positive predictive value and negative predictive value were shown in Table 4. The predictive accuracy value was determined by ROC curve analysis in the validation group. Figure 1 showed that the area under ROC (AUROC) curve in the validation group was 0.851 (0.781–0.921) with statistical significance (*P*<.05). The cutoff value was 2.155. The according sensitivity, specificity, positive predictive value, negative predictive value, and accuracy of the prediction model in the validation group was 76.7%, 77.1%, 80.1%, 72.6%, and 76.9%, respectively.

**Table 4.**
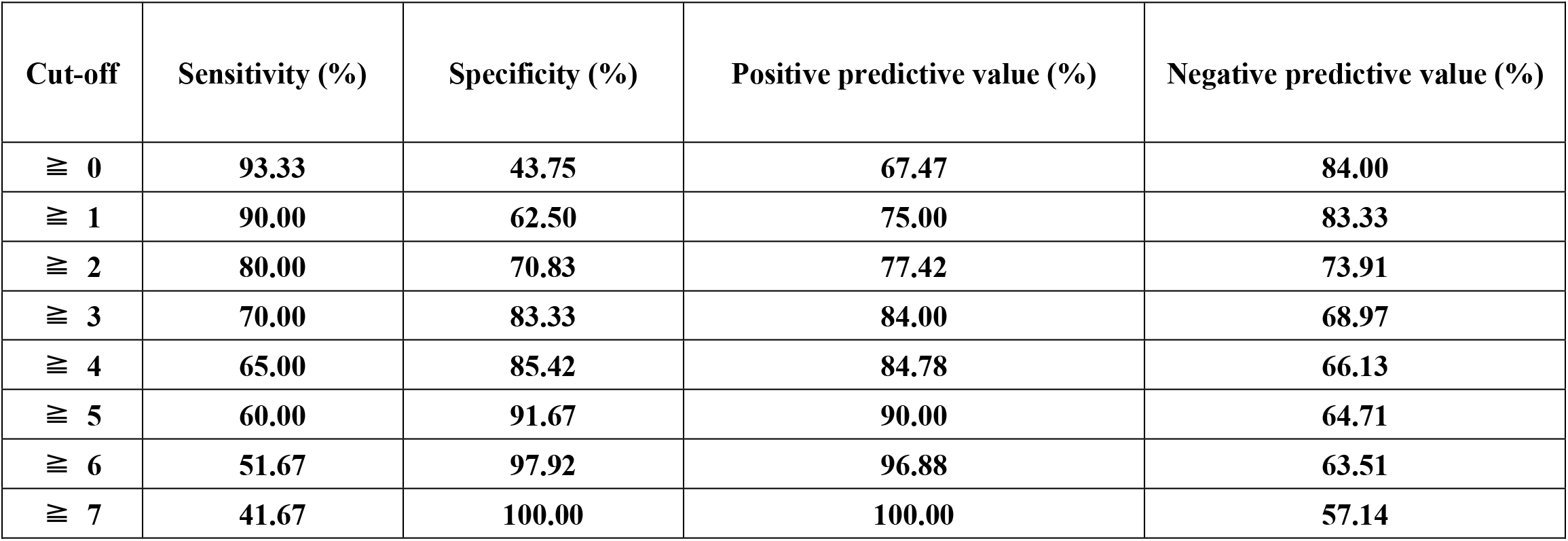
Predicting value of the model for severe pneumonia among DM patients with CAP.

| Cut-off | Sensitivity (%) | Specificity (%) | Positive predictive value (%) | Negative predictive value (%) |
| --- | --- | --- | --- | --- |
| $\geq 0$ | 93.33 | 43.75 | 67.47 | 84.00 |
| $\geq 1$ | 90.00 | 62.50 | 75.00 | 83.33 |
| $\geq 2$ | 80.00 | 70.83 | 77.42 | 73.91 |
| $\geq 3$ | 70.00 | 83.33 | 84.00 | 68.97 |
| $\geq 4$ | 65.00 | 85.42 | 84.78 | 66.13 |
| $\geq 5$ | 60.00 | 91.67 | 90.00 | 64.71 |
| $\geq 6$ | 51.67 | 97.92 | 96.88 | 63.51 |
| $\geq 7$ | 41.67 | 100.00 | 100.00 | 57.14 |

**Figure 1.**
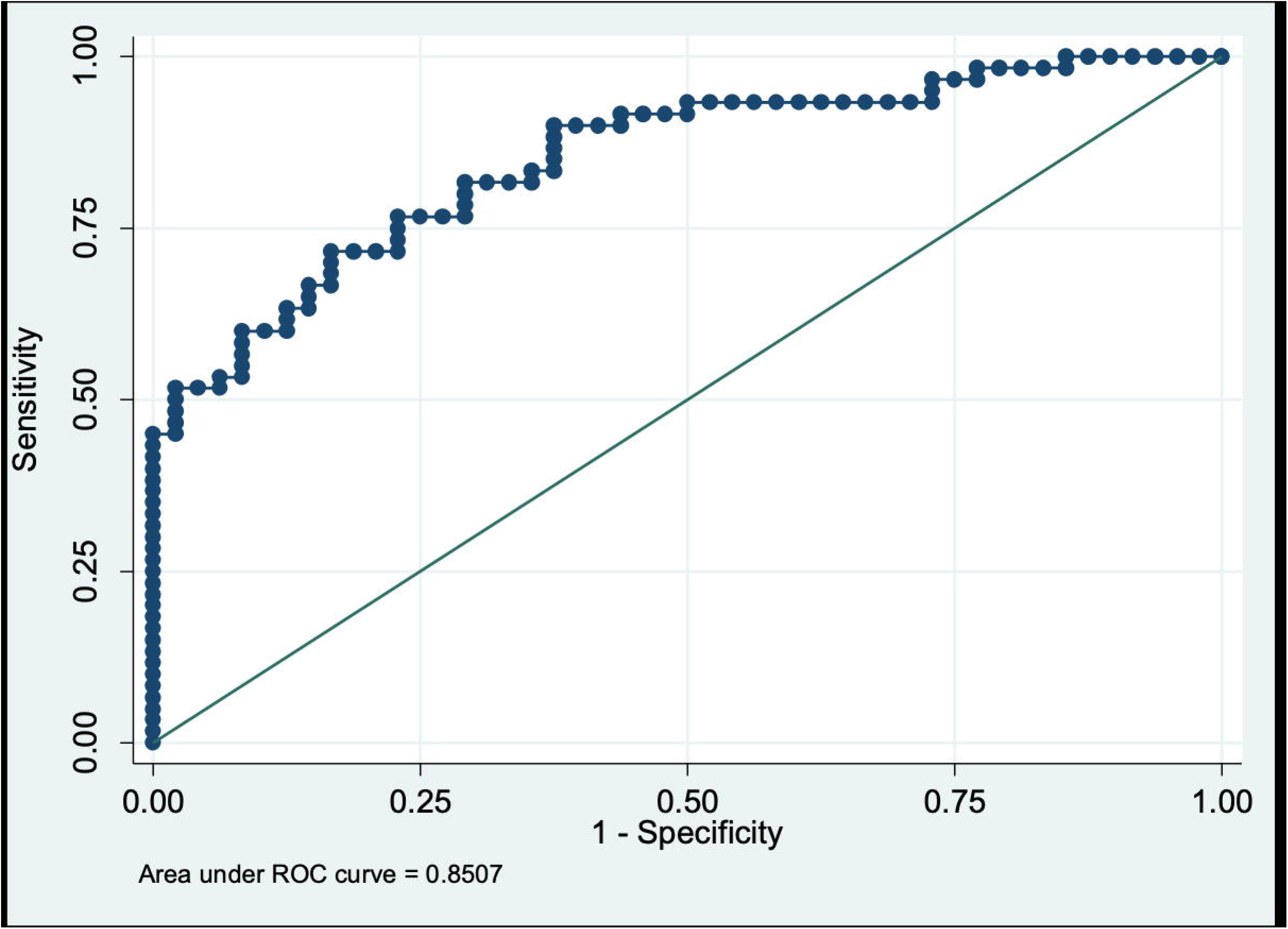
ROC curve analysis of model to predict the progression of CAP among DM patients.

## Discussion

As known, patients with diabetes are considered to have immunocompromised status and are more prone to have chronic organ dysfunction (such as cardiovascular and kidney function). Therefore, diabetic patients have a high risk of developing severe community-acquired pneumonia once diagnosed as community-acquired pneumonia. It is essential to identify these patients early and prevent them from progressing to SCAP to improve clinical outcomes. However, there still lacks useful tools to predict the severity risk of the specific population until now. Therefore, a retrospective study was conducted to develop a tool for predicting severe CAP resulting from CAP among diabetic patients. Our findings suggested that CRP, HbA1C, ALC, and IgM on admission were independently associated with the progression to severe CAP among patients with diabetes and CAP during hospitalization. Furthermore, a combination of these parameters was evaluated and validated for the prediction of severe CAP among diabetic patients and showed high predictive accuracy.

Hyperglycemia is thought to be associated with the immune system and may have a significant effect on the development of SCAP. First, hyperglycemia could reduce the mobilization of polymorphonuclear leukocytes, phagocytic activity, and chemotaxis. Second, some studies found that HbA1c < 8.0% may aid to enhance CD4^+^ T cell response to foreign antigens ^17^. Third, in diabetic patients, glycation of immunoglobulin was correlated with an increased level of HbA1c, which would impair the biological function of antibodies ^18^. In our study, fasting plasma glucose and HbA1C were significantly higher in the severe CAP group than the non-severe CAP group. HbA1C was confirmed an association with the progression of severe CAP by multivariate analysis. Whether hyperglycemia is associated with the mortality and severity of CAP remained unclear. A study conducted by Schuetz et al. evaluated the role of hyperglycemia in CAP patients ^19^. The results suggested that in the non-critical-care setting, initial hyperglycemia was associated with a significant inflammatory response in non-diabetic patients with CAP. However, the association between hyperglycemia at admission and outcome was not found in the diabetic population. It is worthy of being noted that in the diabetic population, persistent hyperglycemia (over 96 h after admission) was associated with adverse clinical outcomes and abnormal levels of CRP. Moreover, patients with diabetes are commonly comorbid with chronic complications (such as cardiovascular disease, renal failure, and pulmonary microangiopathy), which may also influence the progression of CAP and prognosis ^7^. Similarly, a higher proportion of chronic respiratory diseases and kidney diseases were found in severe CAP patients. Moreover, compared with non-severe CAP patients, severe CAP patients have higher levels of CRP, PCT, Scr, BUN, AST, pro-BNP, and lower levels of platelet and albumin, which reflect that severe complications are common in patients with diabetes and severe CAP.

Usually, CAP occurs when the immune system is compromised and encounters a microbial infection. Several variables associated with the immune system have been demonstrated as risk factors of CAP or biomarkers of CAP severity. First, lymphocytes are crucial for immunity. In hospitalized CAP patients, lymphopenia was found as a risk factor of mortality, and lymphocyte count could improve the prediction accuracy of the CURB-65 score for 30-day mortality ^20^. In a study by Bermejo-Martin et al., the association between neutrophil (or lymphocyte) count and 30-day mortality was assessed in hospitalized CAP patients, and the results suggested that patients with lymphocytes <724/mm^3^ had increased mortality by 1.93-fold, irrespective of the CURB-65 score, critical illness, and appropriate antibiotic treatment ^20^. Second, complement factors have been investigated in patients with diabetes. It was reported that compared with patients without diabetes, patients with diabetes have a lower level of C4, which proved to be associated with dysfunction of neutrophils and decreased response to cytokines including interleukin-1 (IL-1), interleukin-6 (IL-6), interleukin-10 (IL-10), interferon-gamma (IFN-γ), and tumor necrosis factor (TNF-α) ^21^. Third, humoral immunity is also essential for the prevention and control of respiratory infections. Impaired humoral immunity would lead to an increased risk for CAP. Torre et al. measured serum total IgG (IgG subclasses, such as IgG1, IgG2, IgG3, and IgG4), IgA, and IgM in CAP patients at the first hospital admission, and the authors found that serum immunoglobulin levels were positively correlated with the severity of CAP and low levels of IgG (or subclasses) were associated with ICU admission and 30-day mortality ^22,23^. Our findings are consistent with these previous studies. In the cohort of patients with diabetes, we found that, compared with non-severe CAP patients, severe CAP patients have lower levels of ALC, CD3^+^ T cell, CD4^+^ T cell, CD8^+^ T cell, IgA, IgM, and IgG. In addition, the progression of CAP to severe CAP was associated with ALC and IgM. In general, it is thought that innate and adaptive immune response may be correlated with the severity of CAP in patients with diabetes, and immunological profiling could provide a valuable tool for severity assessment and prognosis evaluation of CAP among patients with diabetes.

It is vital to develop a prediction tool for early identification of progression to severe pneumonia. In this study, selected combined biomarkers, including CRP, HbA1C, ALC, and IgM, showed a good predictive value of severe CAP among patients with diabetes and CAP. These biomarkers represented an association of severe CAP with inflammatory (CRP), immune status (ALC, IgM), and specific comorbidity factors (HbA1C). Until now, several predictive tools have been characterized in the management of pneumonia. ICU admission is one index that reflects the progression of pneumonia. PSI score has been evaluated in the prediction of hospital/ICU admission. However, the result showed that the PSI perform less well with an AUROC of 0.56–0.85 ^24^. As known, IDSA/ATS has developed new criteria to identify patients with a high risk of ICU admission ^15^. Besides the major criteria mentioned in the guidelines, minor criteria were also suggested for the predictive role. Moreover, in a recent meta-analysis, the prognostic role of minor criteria for ICU admission has been improved, validated, and confirmed ^25^. Also, Kolditz et al. found that the first six minor criteria (without white cell count <4000 cells/mm^3^, platelets <100 000 cells/mm^3^, and hypothermia <36°C) with a combination of hypothermia could predict the death within 7-days ^26^. According to the results ^26^, the minor criteria appear to have excellent negative predictive values for emergency CAP; however, positive predictive values remained very low. Similarly, Renaud et al. evaluated risk factors of ICU admission (within 72 h) in patients with pneumonia and found that hyponatremia, hyperglycemia, leucopenia, and pleural effusion were associated with ICU admission ^27,28^. To assess CAP’s outcome, physicians have evaluated several tools for predicting mortality among high-risk patients. For example, CURB-65 shows a good discriminatory power (AUROC 0.73–0.87) to predict mortality and perform less for predicting ICU admission (0.60–0.78) ^24^. Another study found several risk factors, such as altered mental status, multi-lobar pneumonia and systemic hypotension, age, pneumococcal bacteremia, and discordant empirical antibiotic therapy, have an independent association with mortality within 48 h admission to hospital ^29^.

Of note, we found that a total of 72.1% of patients were diagnosed with pneumonia without definite microbiological etiology in our study (see Figure S1), which is consistent with other studies. The most common etiologies for CAP in DM patients were Klebsiella *pneumoniae* (6.3%), Staphylococcus *aureus* (3.8%), cytomegalovirus (3.8%), mycoplasma *pneumoniae* (3.4%), and influenza virus A (2.9%). We also found that patients with Staphyloccocus aureus or cytomegalovirus more likely progressed to SCAP during the hospital stay. However, we did not include these two pathogens in multivariate analysis for the small sample of microbiological data.

Although several interesting findings were found, our study remains to have several limitations. First, due to the study’s retrospective nature, selection bias may exist, and risk for confounding factors may be indicated. Second, the sample size was relatively small, and further analysis may be required to examine the validity of our findings. Third, we found that more patients with Staphylococcus aureus or cytomegalovirus progressed to severe CAP during hospitalization than those without. However, no significant difference could be found in prediction model group due to the small sample size. Thus, in our further study, we would enroll microbiological etiological data to see if this could predict the progression to SCAP in diabetic patients.

## Conclusion

Our study found that in patients with diabetes and CAP, HbA1C, CRP, ALC, IgM at admission, and their combination could be used to predict the progression to severe CAP during hospitalization with good accuracy. Due to the included biomarkers are common, our finding may be performed well in clinical practice and then improve the early management of diabetic patients with CAP.

## Data Availability

All data referred to in the manuscript are available.

## Ethical Approval

The study protocol was approved by the Ruijin Hospital Ethics Committee, Shanghai Jiaotong University School of Medicine, China. Informed consent from study patients was not required because of the retrospective design of the cohort study.

## Figure Legends

Figure S1. Microbiological findings of CAP patients with DM.

