## Supplemental Figure 1 for "Prediction Model for Severe Community-acquired Pneumonia Development among Patients with Diabetes Mellitus"

### Slide 1
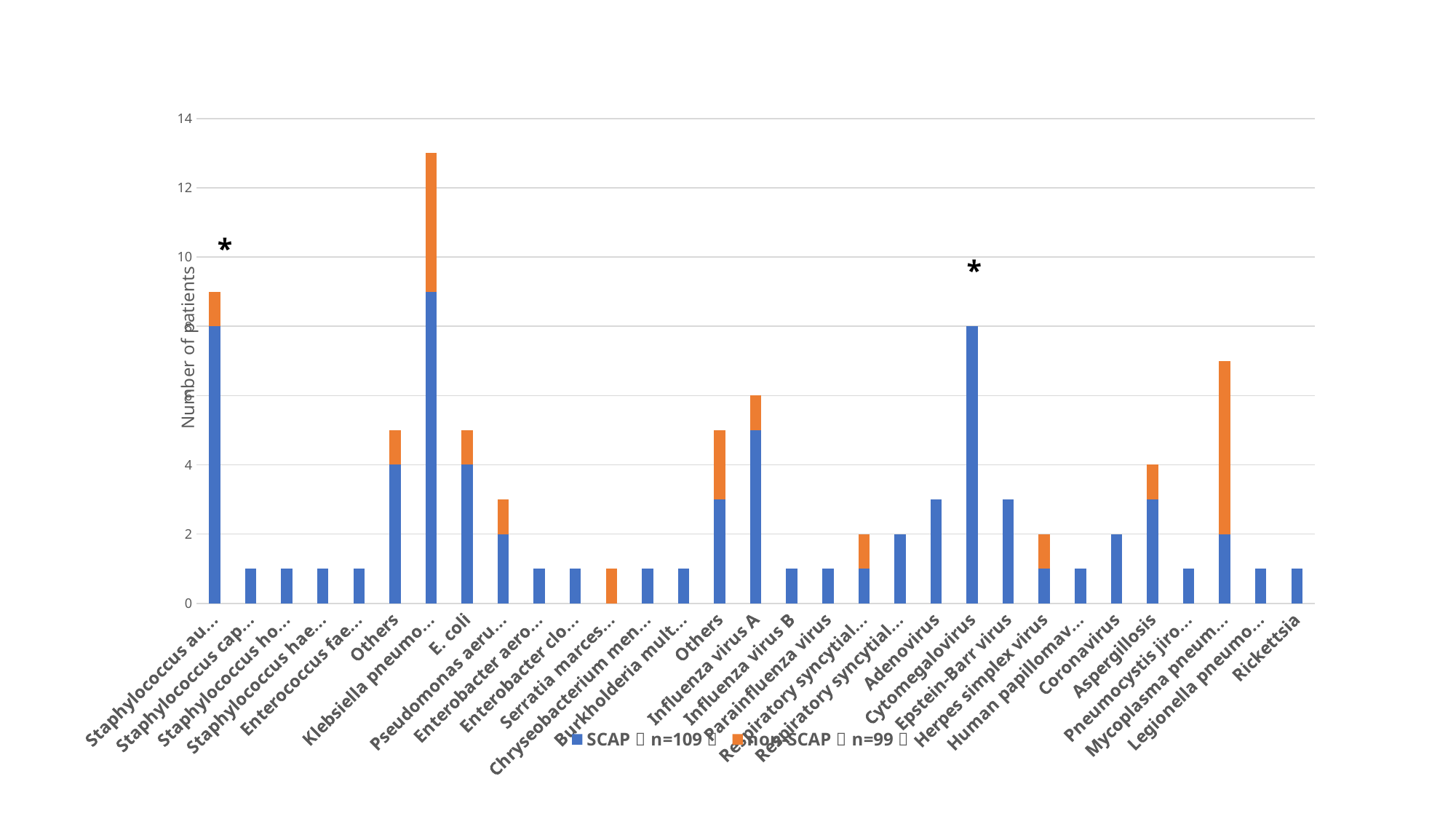

#### Chart
| Category | SCAP（n=109） | non-SCAP（n=99） |
|---|---|---|
| Staphylococcus aureus | 8.0 | 1.0 |
| Staphylococcus capitis | 1.0 | 0.0 |
| Staphylococcus hominis | 1.0 | 0.0 |
| Staphylococcus haemolyticus | 1.0 | 0.0 |
| Enterococcus faecium | 1.0 | 0.0 |
| Others | 4.0 | 1.0 |
| Klebsiella pneumoniae | 9.0 | 4.0 |
| E. coli | 4.0 | 1.0 |
| Pseudomonas aeruginosa | 2.0 | 1.0 |
| Enterobacter aerogenes | 1.0 | 0.0 |
| Enterobacter cloacae | 1.0 | 0.0 |
| Serratia marcescens | 0.0 | 1.0 |
| Chryseobacterium meningosepticum | 1.0 | 0.0 |
| Burkholderia multivorans | 1.0 | 0.0 |
| Others | 3.0 | 2.0 |
| Influenza virus A | 5.0 | 1.0 |
| Influenza virus B | 1.0 | 0.0 |
| Parainfluenza virus | 1.0 | 0.0 |
| Respiratory syncytial virus A | 1.0 | 1.0 |
| Respiratory syncytial virus B | 2.0 | 0.0 |
| Adenovirus | 3.0 | 0.0 |
| Cytomegalovirus | 8.0 | 0.0 |
| Epstein-Barr virus | 3.0 | 0.0 |
| Herpes simplex virus | 1.0 | 1.0 |
| Human papillomavirus | 1.0 | 0.0 |
| Coronavirus | 2.0 | 0.0 |
| Aspergillosis | 3.0 | 1.0 |
| Pneumocystis jiroveci | 1.0 | 0.0 |
| Mycoplasma pneumoniae | 2.0 | 5.0 |
| Legionella pneumophila | 1.0 | 0.0 |
| Rickettsia | 1.0 | 0.0 |*
*
